# Population fraction of Parkinson’s disease attributable to preventable risk factors

**DOI:** 10.1101/2023.05.19.23290231

**Authors:** Haydeh Payami, Gwendolyn Cohen, Charles F Murchison, Timothy R Sampson, David G Standaert, Zachary D Wallen

## Abstract

Parkinson’s disease is the fastest growing neurologic disease with seemingly no means for prevention. Intrinsic risk factors (age, sex, genetics) are inescapable, but environmental factors are not. We studied population attributable fraction and estimated fraction of PD that could be reduced if modifiable risk factors were eliminated. Assessing several known risk factors simultaneously in one study, we demonstrate that all were operative and independent, underscoring etiological heterogeneity within a single population. We investigated repeated blows to head in sports or combat as a potential new risk factor, and found it was associated with two-fold increased risk of PD. Considering modifiable risk factors, 23% of PD cases in females were attributable to pesticides/herbicides exposure, and 30% of PD cases in males was attributable to pesticides/herbicides, Agent Orange/chemical warfare, and repeated blows to the head. Thus, one-in-three cases of PD in males, and one-in-four cases in females could have potentially been prevented.

---

Disability and death due to Parkinson’s disease are rising faster than for any other neurological disease in the world.^1^ In 2016, six million people had PD globally, which was double the number of cases reported in 1990.^1,2^ By 2040, the prevalence is expected to triple to 17 million^2^. These prevalence rates may be underestimated due to lack of awareness of PD and limited access to health care in many populations. The economic burden of PD in 2017 was $51·9 billion in US alone^3^. In 2022 The World Health Organization declared “A pressing need for effective preventive actions … to slow the rising incidence of PD before the burden and costs of treatment overwhelm country health services”^4^.

PD is an etiologically heterogenous syndrome that affects many systems and is progressively debilitating^5,6^. The earliest signs are often gastrointestinal and olfactory dysfunction, depression, and sleep disorder. Movement disorders, the cardinal signs required for diagnosis, appear after decades of prodromal disease and neurodegeneration. By the time of diagnosis, over 80% of dopaminergic cells are already destroyed. At later stages, many develop psychosis and dementia. There is currently no prevention and no treatment to stop neurodegeneration. All clinical trials for disease modifying treatments have failed partly because PD is heterogenous, subtypes are clinically indistinguishable, and biomarkers to select appropriate patient population for trials are lacking. PD is likely not a single disease, rather a syndrome with a range of causes, all converging to produce the phenotype we call PD^5,7^. Three to five percent of PD are caused by pathogenic mutations in any one of causal genes, which are rare, and have seemingly different functions, e.g., *SNCA, PRKN, LRRK2, PINK1*^6^. The vast majority of PD remains idiopathic, thought to be caused by environmental triggers interacting with intrinsic risk factors such as genetic susceptibility^6,8^.

Currently, the most compelling weapon against the global rise in PD is elimination of risk factors^9^. Intrinsic risk factors, which are not readily amenable to prevention, include increasing age, the male sex, positive family history of PD, and genetic causes and susceptibility variants^6,10-15^. Among environmental factors, neurotoxicant exposure, especially pesticide and herbicides, has been robustly associated with increased risk of PD^8,16-18^. Head injury is also common prior to development of PD but its causal relationship is questionable^19-22^. Cigarette smoking and coffee drinking are associated with reduced risk of PD^23,24^, and their protective effects are independent and additive such that non-smokers and caffeine users have extremely low risks^25,26^.

Here, we investigated several factors simultaneously in a single population. Doing so enabled us to compare their effects side-by-side, test inter-dependence and overlap among them, and assess the burden of disease due to modifiable risk factors. We investigated family history of PD, exposure to pesticides/herbicides, Agent Orange (an herbicide used in the Vietnam war)/chemical warfare, mild or moderate traumatic brain injury (MTBI)/concussion, and repeated blows to the head in sports or military as a potential new risk factor. We addressed the following questions. (1) Are all risk factor operative in one population? If only one or few risk factors predominate in a population, that would be an opening to teasing out etiologic heterogeneity. (2) Do repetitive hits to the head that do not have an immediate clinical manifestation increase risk of PD? There is growing evidence linking repetitive hits to the head in collision sports to neurodegenerative disease later in life^22,27^. Prior PD studies have defined head injury as hospital-treated MTBI or concussion^19-22^. It is not known if repetitive hits to the head with no immediate signs of injury increase PD risk. (3) Are the risk factors independent of each other? The few studies that have addressed interdependence among protective and risk factors found them to be independent^25,26,28^, and suggested the risk factors mark etiological subtypes of PD^28^. (4) What percentage of PD could potentially be reduced if modifiable risk factors were eliminated? Population attributable fraction (PAF) is an epidemiological metric commonly used to estimate the fraction of disease that is attributed to a risk factor^29,30^. It has been applied widely to many disorders including dementia^31^, but not to PD. One study has calculated population attributable risk (PAR not PAF) for PD reporting that 54% of PD risk is explained by the combined effects of family history of PD, not smoking, and occupational exposure to lead, copper, and insecticides^32^. Here we calculate PAF, with intervention as the goal, for potentially modifiable risk factors. To our knowledge, this is the first study of PAF for PD.

The study was conducted in Birmingham, Alabama, in the center of the Deep South of the United States. The Deep South is understudied. Little is known about epidemiology of PD in the Deep South except that in surveys of United States, the Deep South states were found to have high incidence of PD^10^, and high rates of mortality from PD^33^. We show that all risk factors including repeated blows to the head, but possibly not MTBI/concussion, are operative in this geographically and culturally defined population, the risk factors are independent, and some are actionable candidates for disease prevention, and plausible markers for disease subtyping to reduce heterogeneity in clinical trials.

## Results

### Subject characteristics and PD features

Study included 808 persons with PD (PwP) and 415 neurologically healthy controls (NHC). Nearly all study participants were living in the Deep South and peripheral states and 77% of them were born in the Deep South. This relatively homogenous population, compared to metropolitan regions of the world, provided a convenient sample to determine which features and risk factors are operative in a single population. All known features of PD that were tested were captured in this population with statistical significance (**Table 1**). Notably, among PwP, 63% were males (P<1E-5). Constipation was five-fold more prevalent in PwP than in NHC (P=4E-25). Twice as many PwP reported having lost >10 pounds in the prior year than NHC (P=4E-7). Ten percent of PwP vs. none of the NHC reported having had rapid eye movement sleep behavior disorder (RBD), a rare disorder that often leads to PD (P=2E-6). Recapitulating these features establishes robustness of the data.

**Table 1.** Enrollment, subject characteristics, and features of PD.

| Subject Characteristics | PD |  | NHC |  | PD features |  |
| --- | --- | --- | --- | --- | --- | --- |
|  | N with data | Summary statistics | N with data | Summary statistics | OR[95%CI] | P |
| Sample size | 808 | - | 415 | - | - | - |
| Age | 808 | 68.3±8.7 | 415 | 66.1±8.6 | - | - |
| PD age at onset | 808 | 59.3±11.0 | - | - | - | - |
| Jewish ancestry | 597 | 16 (3%) | 313 | 9 (3%) | - | - |
| Black or African American | 804 | 20 (2%) | 410 | 6 (1%) | - | - |
| Hispanic or Latino | 787 | 13 (2%) | 403 | 3 (0.7%) | - | - |
| White | 804 | 772 (96%) | 410 | 398 (97%) | - | - |
| Residents of Deep South | 799 | 764 (96%) | 403 | 376 (93%) | - | - |
| Residents of DS Periphery | 799 | 32 (4%) | 403 | 26 (6%) | - | - |
| **PD Features** |  |  |  |  |  |  |
| Sex (N & % male) | 808 | 512 (63%) | 415 | 125 (30%) | - | <1E-5 |
| Constipation | 775 | 345 (45%) | 402 | 57 (14%) | 4.8 [3.5-6.7] | 4E-25 |
| RBD | 479 | 47 (10%) | 229 | 0 (0%) | 50.4 [3.1-821.5] | 2E-6 |
| Weight loss | 790 | 212 (27%) | 406 | 56 (14%) | 2.3 [1.7-3.1] | 4E-7 |

A total of 1466 individuals were enrolled, of whom 1223 completed the questionnaires and were used in the present study, including 808 persons with PD and 415 neurologically healthy controls (NHC). Characteristics are shown with the number of individuals for whom specified data were available and analyzed (N with data). Summary statistics is the mean and standard deviation for quantitative traits, and the number of individuals and % of total who are positive for the characteristic for dichotomous traits. Deep South (DS) includes the states of Alabama, Mississippi, Georgia, South Carolina, and Louisiana. DS periphery states are Texas, Tennessee, North Carolina, and Florida. Question on REM sleep behavior disorder (RBD) was added at the start of the second wave of data collection and was not asked of 316 PD and 181 NHC of the first wave. To assess robustness of data, features that are known to be more prevalent in PD were tested. Preponderance of male sex among PD was tested as proportion of persons with PD who were male (63%) vs. expected for null hypothesis of no sex difference (50%), using Z test (one proportion test) and showing the significance (P). Constipation, RBD and weight loss were tested comparing PD to NHC, using odds ratio (OR), 95% confidence interval (CI) of OR, and significance (P) denoting the difference between PD and NHC.

### Risk factors

We collected data and tested association of the following risk factors with PD simultaneously in a single study population of 808 PwP and 415 NHC: family history of PD, exposure to pesticides/herbicides, exposure to Agent Orange/warfare chemicals, and MTBI/concussion. We also investigated repeated blows to the head in sports or combat as a potential risk factor. The exact questions that were used to collect the data are shown in **Table 2**.

**Table 2.**
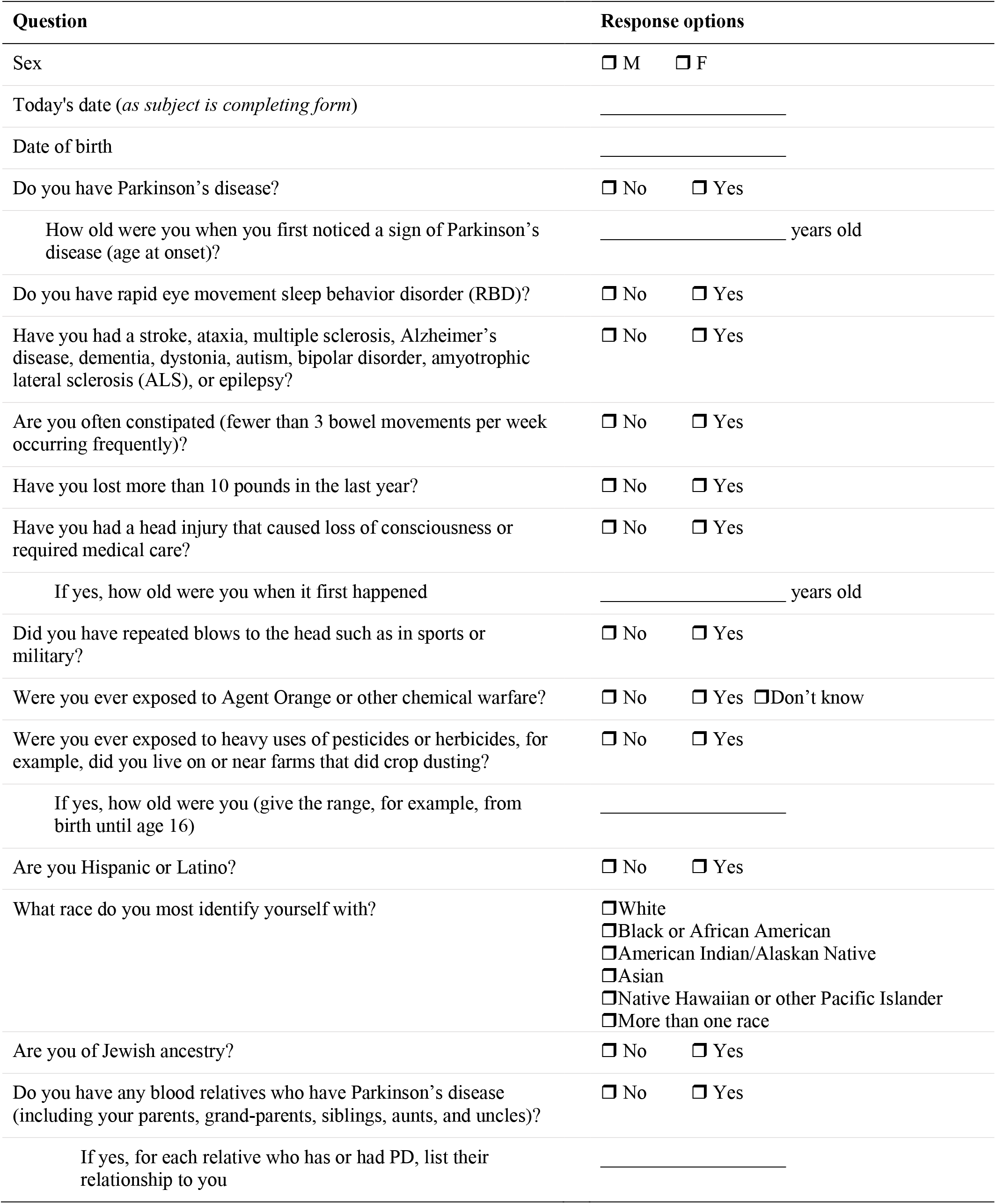
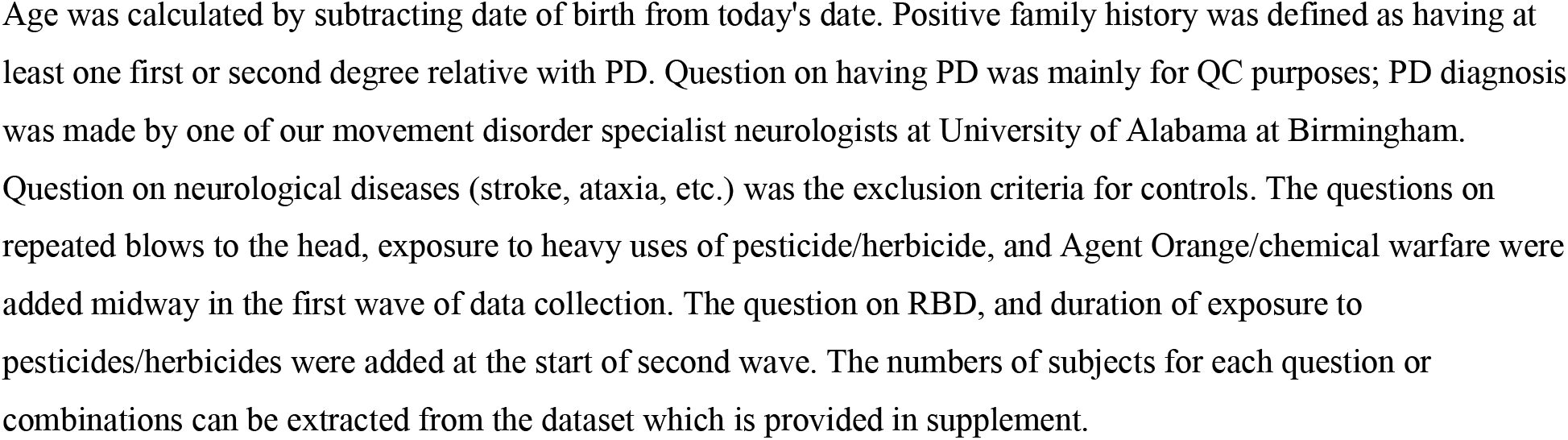
Questions used to collect the data.

We detected a strong and robust association signal for every risk factor examined except MTBI/concussion. **Table 3** shows raw unadjusted odds ratios (OR), as if each risk factor was assessed individually, first for all subjects, and then stratified by sex. **Table 4** shows results of conditional analysis where we included all risk factors in one model and estimated adjusted OR and P values for each risk factor conditioned on all other risk factors including age. Sex could not be included in conditional analysis because of collinearity of male sex with repeated blows to the head (all male) and exposure to Agent Orange/warfare chemicals (all male except one female).

**Table 3.**
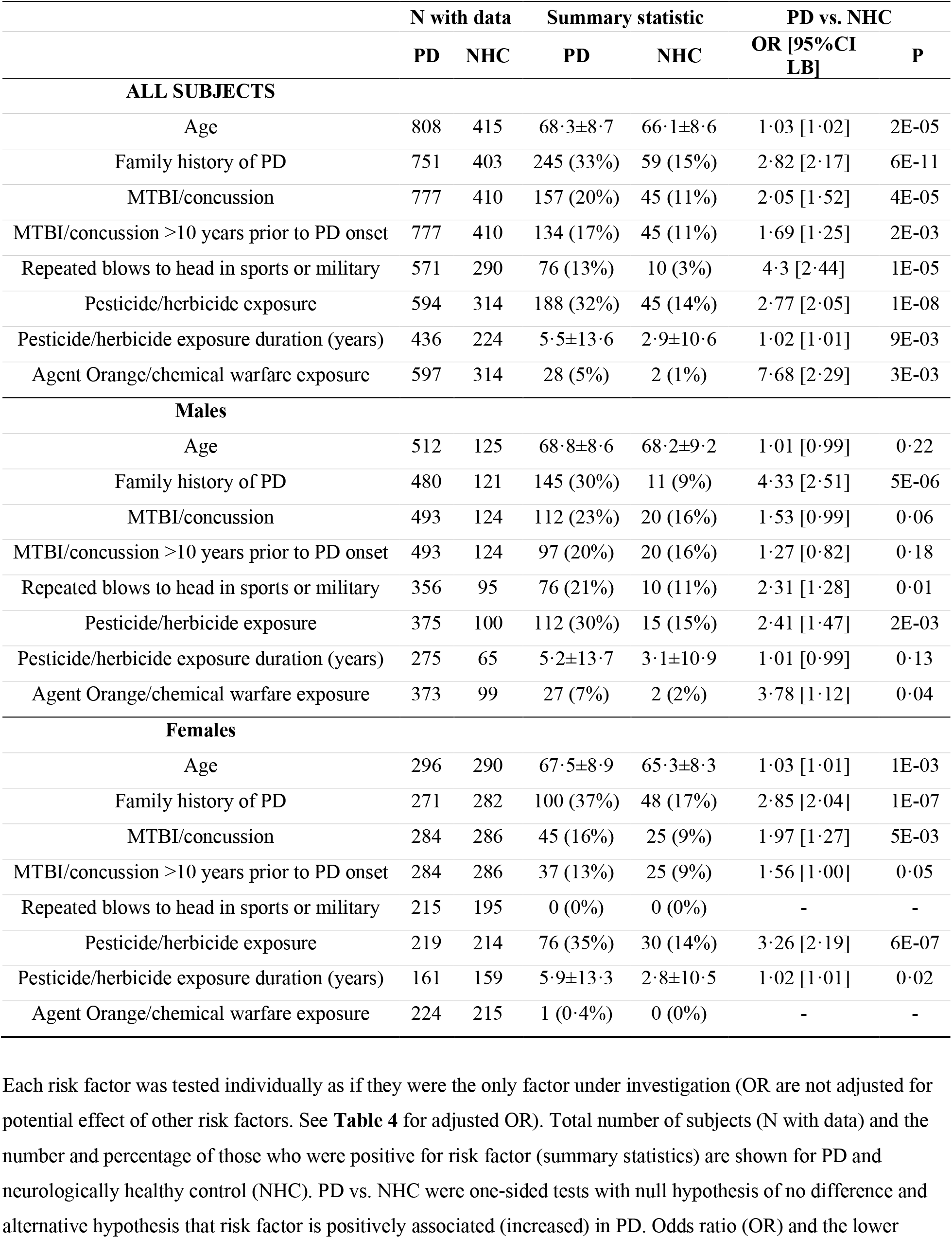
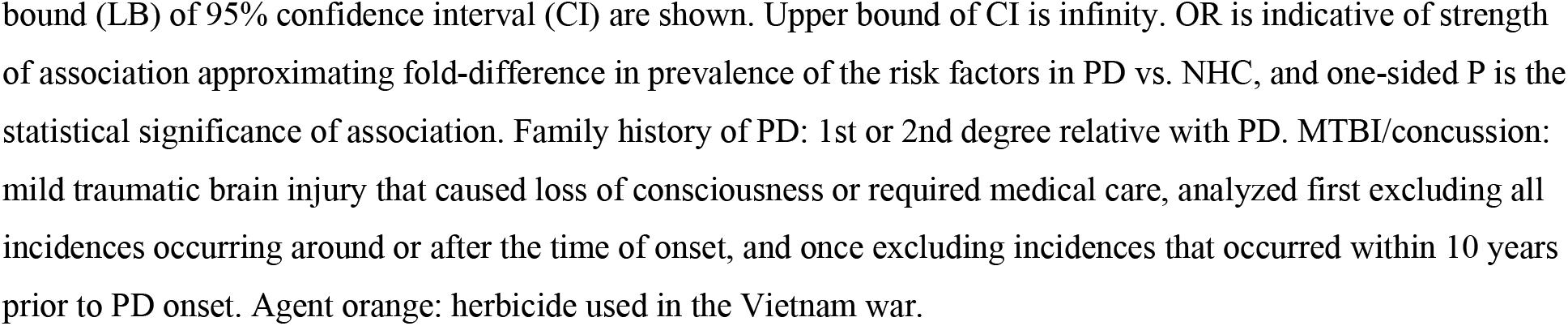
Assessing association of risk factors with PD in a single population.

**Table 4.**
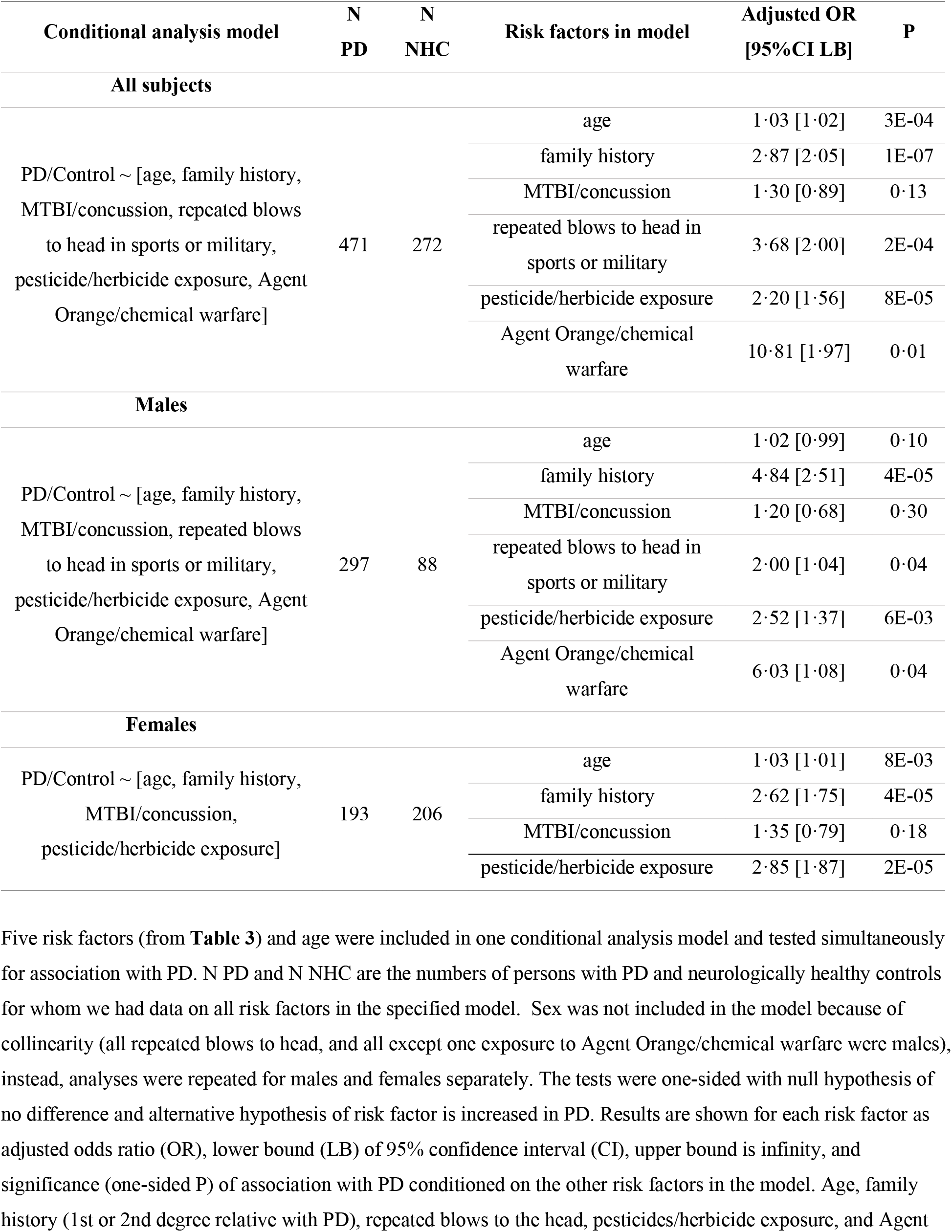
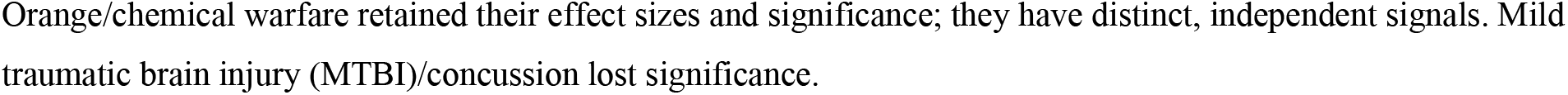
PD Risk factors are independent of each other.

**Table 5.** Population attributable fraction (PAF) of preventable risk factors.

| Risk factor | Group | PAF [95%CI] | Determinants of PAF |  |
| --- | --- | --- | --- | --- |
|  |  |  | adjusted OR | Prevalence in PwP |
| PAF for Individual risk factors |  |  |  |  |
| Pesticide/herbicide exposure | All | 16% [9%-24%] | 2.20 | 30% |
| Pesticide/herbicide exposure | Females | 23% [12%-34%] | 2.85 | 35% |
| Pesticide/herbicide exposure | Males | 17% [7%-27%] | 2.52 | 28% |
| Agent Orange/chemical warfare | Males | 6% [0.03%-12%] | 6.03 | 7% |
| Repeated blows to head in sports or military | Males | 10% [0.3%-20%] | 2.00 | 20% |
| Joint PAF for combination of risk factors |  |  |  |  |
| Repeated blows to head in sports or military, pesticide/herbicide & Agent Orange/chemical warfare | Males | 30% [7%-49%] | - | 43% |

Family history of PD was three-to four-times more common in PwP than NHC and was highly significant and robust to sex-stratification (**Table 3**), adjustments in conditional analysis, and sex-stratified conditional analysis (**Table 4**).

Pesticide/herbicide exposure was associated with a robust two-to three-fold increased risk of PD, in all subjects, in both sexes (**Table 3**), and when adjusted for other risk factors (**Table 4**). Duration of exposure was also significantly longer, almost double, for PwP than NHC (**Table 3**).

Unadjusted result for MTBI/concussion was initially significant showing a two-fold increase in PD (**Table 3**). However, the signal weakened in sex-stratified data (**Table 3**) and was further reduced and lost significance in conditional analysis (**Table 4**). Prior studies have questioned the causal relationship of MTBI/concussion with PD because it is possible that balance problems in prodromal PD may be causing falls and head injury (reverse causality). One study showed that association of MTBI/concussion with PD was limited to the events occurring within 10 years of PD diagnosis^20^. In our study also, excluding MTBI/concussion within 10 years of PD onset, had a notable effect on weakening the signal (**Table 3**).

The unadjusted results for Agent Orange/chemical warfare and repeated blows to the head are skewed. Note that (a) there is a sex differential in this study, caused initially by inevitable increased prevalence of PD in males, and exaggerated by preferential selection of spouses as controls, and (b) sex could not be included in the conditional models because of strong collinearity of male-sex with repeated blows to head and Agent Orange/chemical warfare. Hence, the sex-stratified analyses are critical for interpreting the results.

Repeated blows to the head was initially estimated as four-fold more common in PwP than NHC (**Table 3**). However, repeated blows to the head was reported by males only, and in males, it was associated with two-fold increased risk and was significant both individually (**Table 3**) and after adjusting for other risk factors including MTBI/concussion (**Table 4**).

Exposure to Agent Orange/chemical warfare yielded an unadjusted eight-fold increased risk (**Table 3**). Only one female, who had PD, reported exposure to Agent Orange/chemical warfare, the rest of the exposed were males. In males, Agent Orange/chemical warfare yielded an unadjusted four-fold increased risk. Interestingly, Agent Orange/chemical warfare association gained strength in conditional analysis, yielding a 10-fold increase in all subjects, and six-fold increase in males (**Table 4**).

We conclude that family history, exposure to pesticides/herbicides, Agent Orange/chemical warfare (as seen predominantly in males), and repeated blows to the head (as seen only in males) are all operative in this population, and that they have distinct independent effects on PD risk. Demonstration of independence does not preclude possibility of interaction, for example, between a certain genetic locus (captured by family history) and a chemical compound (captured by pesticides/herbicides). It does however establish each risk factor as a significant contributor to risk, independent of other factors.

### PAF

PAF is an epidemiological metric to assess public health impact of a risk factor. PAF incorporates the strength of the risk (OR) with prevalence of risk factor to provide an estimate for the percentage of disease that would be reduced if risk factor were eliminated. We calculated PAF for risk factors that are modifiable, in the appropriate groups i.e., pesticide/herbicide exposure in males and females, Agent Orange/chemical warfare in males, and repeated blows to head in males. In calculating PAF we used adjusted OR (from **Table 4**) that were conditioned on all risk factors including age, family history, MTBI/concussion, pesticide/herbicide exposure, Agent Orange/chemical warfare, and repeated blows to head, as applicable. Hence, all PAF estimates are adjusted PAFs. Adjusted PAF for pesticide/herbicide exposure was 23% [12%-33%] for females and 17% [7%-27%] for males. In addition, among males, adjusted PAF was 10% [0·3%-20%] for repeated blows to the head, and 6% [0·03%-12%] for Agent Orange/chemical warfare exposure.

In women, only one modifiable risk factor was detected. In men, there were three. To assess fraction of disease in males that was attributed to all three risk factors, we calculated joint PAF for the combination of pesticide/herbicide exposure, Agent Orange/chemical warfare, and repeated blows to head. Joint PAF assumes that risk factors are independent. In conditional analysis (**Table 4**) we established that each risk factor has an independent effect on PD risk. To assess if there is interaction (e.g., synergism), we tested pairwise interaction for the three risk factors in males. There was no evidence for interaction in this dataset. The interaction P values were P=0·99 for [pesticide/herbicide exposure* Agent Orange/chemical warfare], P=0·99 for [Agent Orange/chemical warfare*repeated blows], and P=0·56 for [pesticide/herbicide exposure* repeated blows to head]. Having established independence and lack of interaction, we calculated joint PAF for Agent Orange, pesticide/herbicide exposure, and repeated blows to head in sports or military in males as 30% [7%-49%].

## Discussion

### Summary

We investigated multiple risk factors simultaneously in a single population and addressed four questions: (1) Are all major risk factor of PD operative in one population? We had hoped that if only one or few risk factors predominate in a population, that would be an opening to teasing out etiologic heterogeneity. Nearly all features and risk factors tested were associated with PD. Thus, the extensive heterogeneity of PD is encapsulated even in this relatively isolated population in the Deep South. (2) Do repetitive hits to the head that do not have an immediate clinical manifestation increase risk of PD? Repeated blows to the head, which is common in collision sports and do not normally require medical care, was associated with two-fold increases risk of PD. (3) Are the risk factors independent of each other? Family history, toxicant exposure, and repeated blows to the head are independent risk factors with little overlap, which is consistent with a prior report^28^. (4) What percentage of PD could potentially be reduced if modifiable risk factors were eliminated? One in three cases of PD in males, and one in four cases in females were attributed to modifiable risk factors and could have potentially been prevented. Results have two broad implications. First is prevention. PD is a complex disease with both intrinsic (genetics, aging) and extrinsic (environmental, life-choices) risk factors. While intrinsic factors are unescapable, extrinsic triggers may be preventable, thereby averting the disease. PAF estimates suggest a substantial fraction of PD is potentially preventable. Second is potential markers for disease subtypes. Clinical trials for neuroprotective treatments have all failed, due in part to lack of biomarkers to distinguish subtypes of PD. That the risk factors for PD are independent indicates family history of PD, repeated blows to head, and pesticide/herbicide exposure tag etiologic subtypes of PD.

### PAF

PAF bridges science and decision making. It provides an estimate of excess risk that could potentially be eliminated, a starting point for discussing intervention. For PAF to be meaningful, several requirements must be met. Intervention must be possible. To that point, we calculated PAF only for modifiable risk factors and not for age, family history, or sex. The term “attributable” has a causal interpretation. The risk factors selected for PAF in this study have been causally linked to PD^34-36^. There is probably variation in genetic susceptibility that make people more or less vulnerable to potential damage caused by toxicant exposure or head injury. The cause is technically gene*environment interaction. However, for PAF, the environmental trigger meets the definition/assumption of causality. Another requirement for accurate estimation of PAF is no bias in the study design and data being valid and accurate. We assert the robustness of our data from the strict QC measures that were implemented, subject selection was sequential and from only one movement disorder clinic, methods were standardized, and the resulting data recapitulated every known feature and risk factor of PD with high statistical significance and reasonable effect size. ORs were adjusted for all risk factors in the study, and adjusted PAFs were calculated using the adjusted ORs. Since risk factors are not mutually exclusive (a farmer exposed to toxicants could also have played a collision sport and had repeated blows to the head) the overlaps were accounted for by correct calculation of joint PAF^30,37^, having first established independence and absence of interaction, and then using adjusted ORs that were conditioned on all other risk factors. Finally, the interpretation of PAF is dependent on how exactly the exposure was defined; to that end, we have provided the exact questions that were asked to solicit the data in **Table 2**. In short, we believe we have met the assumptions of PAF, conducted a rigorous study, and results are robust, suggesting a substantial fraction of PD could have been prevented if these individuals had not been exposed to heavy uses of pesticide/herbicides or Agent Orange/chemical warfare and had avoided activities that caused repeated blows to the head. We note that this is the first attempt at estimating PAF for PD and point out that the PAF estimates will be population specific depending on the distribution and frequency of risk factors.

### Pesticides/herbicide

Pesticides and herbicides are among the most robust risk factors for PD^8,16-18^. Some pesticide and herbicides are inherently neurotoxic^38^. For instance, particular insecticides have been designed and developed to target neuronal pathways that are shared between insects and animals. PAF estimates obtained here suggest that exposure to pesticides/herbicide accounted for 23% of PD cases among females and 17% among males. This is an underestimate because our calculations were based on self-reported exposures to heavy uses of pesticides/herbicides and did not include exposure through contaminated soil, water, and consuming contaminated fruits, vegetables, meat, and fish^39-41^. US allows use of 72 pesticides that are banned in EU, and their usage in US has stayed constant or increased in the past decade^42^. Pesticide levels in surface waters are higher in US than EU and China^43^. Given such a robust risk and prevalence, and the potential for disease prevention, it is prudent that steps be taken to regulate access to and limit exposure to these toxicants, both acutely and chronically.

Our study does not identify any one particular chemical that is directly associated with PD. In the US, few states document the use of pesticides and herbicides, with very few making such data publicly accessible. Thus, it is difficult to pinpoint the risk effect of one specific chemical, outside of few locations (such as California^44^). Mandatory and enforced registration of pesticide application would be a first step to better understand exactly which chemicals we are exposed to, at what dose, and for how long. We still depend on certain pesticides and herbicides for modern agriculture and human health. Pyrethroids, for example, are important tools to control mosquito populations for limiting malaria, West Nile and Dengue viruses^38^. However, for certain chemicals with robust evidence of long-term harm, such as paraquat that has been firmly linked to PD in both human and experimental settings^5,38^, complete bans, as done in other countries, should seriously be considered.

### Head injury

The association of PD with repeated blows to the head that do not cause concussion or require medical care was a notable finding in this study. Prior PD studies defined head injury as MTBI that caused concussion or needed medical care. MTBI/concussion is often found to be more common among PwP, however, causality has been questionable^19,20^. Is it MTBI/concussion that increases risk of PD or is it impaired balance in prodromal PD that increases risk of falls and head injuries? MTBI/concussion was more common in PwP in our study as well, but it lost its effect and significance when adjusted for other risk factors. We also saw a weakening signal when we excluded MTB/concussion within 10 years prior to PD onset. MTBI/concussion was the only risk factor that did not retain its signal after adjusting for other risk factors, even when all injuries up to onset of PD were included. Repeated blows to the head, however, had a robust signal of association that withstood adjustment for all other risk factors including MTBI/concussion. The most common source of repeated blows to the head is collision sports. Studies have shown adverse consequences of collision sports (mainly American football and soccer) on brain health later in life^22,27,45^. It is well-publicized in the popular media that football players go on to suffer high rates of depression, mood and behavioral problems, cognitive loss, dementia, and chronic traumatic encephalopathy. Despite high-profile national coverage by media on the dangers, American football remains highly popular in the US. In our study population, only men reported having had repeated blows to the head as youths. The popularity of collision sports is not waning among boys and is rising among school age girls. It is therefore imperative to implement effective safety measures for football, soccer, and all collision sports.

### Caveats, Limitations, Confounders

Repeated blows to the head as a risk factor for PD needs to be tested and confirmed in other populations. PAF estimates will vary across populations depending on the prevalence of the risk factors.

The PAF estimates reported here were based on a population of older adults and their past experiences that may have put them at risk. They are not necessarily predictive of the future. For example, in this cohort of older Southerners, no female reported repeated blows to the head due to sports or military, and only one female (with PD) had been exposed to Agent Orange. In the 1900’s when these women were young, women did not partake in collision sports or military, as they do now. The PAFs for future will change, for the better or worse, depending on the actions we take now to clean our environment and improve health and safety standards.

As noted in sections above, we incorporated guards against potential confounders in our study design and statistical analyses. Here, we address two other commonly cited potential confounders in epidemiological studies of PD: reverse causality, and risk aversion. MTBI/concussion, although it was eliminated statistically, is a good example of confounding by reverse causation. Difficulty with balance may be present in prodromal PD and cause falls resulting in head injury, hence the possibility of PD causing head injuries, not vice versa. Reverse causality is not likely for repeated blows to head because they occur many decades prior to onset of PD, in children and young adults who partake in collision sports or military. Risk aversion, noted to be a premorbid personality trait^46^, complicates interpretation of negative associations such as smoking with PD. For example, while nicotine is known to be neuroprotective, it is still unclear if smoking is protective against PD or individuals who will eventually develop PD are averse to smoking due to its associated risks. This argument does not apply equally to risk factors as it does to protective factors. If anything, risk averse individuals would avoid collision sports, military, and toxicant exposure. Thus, if risk avoidance is a confounder for association of PD with repeated blows to the head, or with toxicant exposure, their true effect sizes could be larger than our estimates.

### Disease subtypes

PD is a heterogenous syndrome with more than one cause^5,6^. Currently, the subtypes within idiopathic PD are indistinguishable. Disease heterogeneity, and unavailability of biomarkers to distinguish disease subtypes, is considered the main reason that clinical trials for disease modifying PD drugs fail. It has been proposed that the major risk factors for PD, that are independent of each other in their association with PD, represent markers for etiological subtypes^28^. Our study was set in the Deep South, which is a culturally unique and relatively isolated and stable population in the US. Over 95% of Subjects in this study live in the Deep South and 77% of them were born here. The relative uniformity in this population compared to more cosmopolitan regions of the world provided a desirable setting to test all risk factors simultaneously, in search of population-specific risk factors. We found all risk factors and features of PD robustly associated with PD in our study population. Our results indicate risk heterogeneity is present, to full extent, within even a relatively homogenous population.

Extensive efforts and cost are being spent in search of molecular biomarkers of disease subtypes in blood, cerebral spinal fluid, and microbiome. In a breakthrough, alpha-synuclein seed amplification assay (SAA) was recently described as a diagnostic biomarker in symptomatic PD and some high-risk populations^47^.SAA’s detection power varies by genetics and clinical features, which underscores the etiological heterogeneity of PD, although SAA does not distinguish disease subtypes. Currently, the only markers that can distinguish some PD subtypes are the highly penetrant pathogenic mutation in a causal gene (*SNCA, PRKN, LRRK2*, etc.), which are rare. For common idiopathic PD, there is currently no clear way to define and divide the subtypes^48^. As efforts for molecular biomarker discovery progress, we propose a simple, crude but potentially powerful alternative for the meantime. Here, we demonstrated that repeated blows to the head, exposure to pesticides/herbicides, and having a family history of PD are each an independent risk factor for PD. They all precede PD. Each is a biologically plausible cause. Their independence suggests they could represent markers for different etiological subtypes of PD. Belvisi et al.^28^ also found family history of PD and exposure to pesticides (and metals and anesthesia) to be independent risk factors, and surmised that they mark etiological subtypes of PD. We agree. To that end, we propose that taking a brief history – as simple as the questions in **Table 2** - could help gauge and potentially reduce heterogeneity in clinical trials and improve odds of success.

## Methods

### Enrollment of Study participants

We have complied with all relevant ethical regulations. The study was approved by the Institutional Review Board (IRB) for Protection of Human subjects at the University of Alabama at Birmingham (UAB) and by the Office of Human Research Oversight (OHRO) of United States Department of Defense (DoD, funding agency). All subjects signed informed consent. No compensation was provided for participating in the study.

We enrolled 981 persons with PD (PwP) and 485 neurologically healthy controls (NHC). Inclusion criteria for PwP were diagnosis of PD by a movement disorder specialist neurologist at UAB according to UK Brain Bank criteria^49^, and informed consent. PwP were enrolled sequentially as they were seen at clinic. Controls consisted of patients’ spouses and community volunteers. Inclusion criteria for controls were informed consent and self-report of not having PD, stroke, ataxia, multiple sclerosis, Alzheimer’s disease, dementia, dystonia, autism, bipolar disorder, amyotrophic lateral sclerosis, or epilepsy (hence, neurologically healthy controls, NHC).

Enrolment occurred in two waves: July 2015-July 2017 (N=316 PwP and 181 NHC) and October 2018-March 2020 (N=665 PwP and 304 NHC). The sample size was determined by the arrival of COVID19 pandemic in the Deep South, when we stopped enrolling.

### Data collection

Subjects were asked to fill out two questionnaires. The Environmental and Family History Questionnaire (EFQ) was first developed 1994 for the NeuroGenetics Research Consortium (NGRC), and has been published for three decades^12,50^. The Gut Microbiome Questionnaire (GMQ, the source for gastrointestinal data) was developed in 2014 and has also been published^50^. Questionnaires were self-administered with no interference from research staff. In the first enrollment wave, subjects completed the questionnaires in the clinic. In the second wave, subjects took the questionnaires home, and 74% of PwP (N=492) and 77% of NHC (N=234) completed and returned them. The final analytic sample size with completed questionnaires was 808 PwP and 415 NHC (**Table 1**).

Questions that were asked to collect the data used in this analysis are shown in **Table 2**. Positive family history was defined as having at least one first-or second-degree relative with PD; negative family history was absence of PD in first- and second-degree relatives; and indication of family history without specifying the degree was classified as unknown^12,50^. MTBI/concussions events (N=19) and exposures to pesticides/herbicides (N=5) which occurred in the same year or after onset of PD were not counted as events. When limiting MTBI/concussions to incidences that occurred more than 10 years prior to PD onset, the incidences within 10 years were treated as no event.

### Quality control

QC checkpoints were implemented at every step possible during data collection, data entry and data analysis. Briefly, questionnaires were reviewed at arrival for inconsistency across forms, missing data, and potential mix-up at home of case-control couples. Issues were resolved by contacting the subjects or reviewing medical records, otherwise the datapoint was excluded from analysis. All data (100%) were entered in computer twice, once in a Progeny database (**Supplementary Material/Software)**, and once in Microsoft Excel, then downloaded and compared, data entry errors identified and corrected. Progeny was used as the main database. When subjects provided a non-standard answer (e.g., “teenager” instead of specifying age), standard rules were created and applied uniformly to all such answers for all subjects (ex. teenager=16). When in doubt, data were recorded as “Unknown” and made distinct from “No”. Statistical analyses were conducted by different analysts independently, one using manual calculation and one using a code and software pipeline, and were cross-checked for accuracy. Here, we present the results obtained via pipeline and the accompanying code.

### Statistics

The number of subjects with data for each item can be extracted from the **Supplementary Material/Data** and are also given for each test in the Tables. For software, the URLs and version used are given in **Supplementary Material/Software**. The code for data analysis is available on Zenodo [link will be provided in peer reviewed paper].

Differences in frequencies of features (constipation, RBD, weight loss) in PwP vs. NHC were tested using Yates continuity-corrected OR, using the function ‘Prop.or’ from the pairwiseCI R package specifying “CImethod=‘Woolf”.

Association between risk factors and disease was tested under the null hypothesis of no difference and the alternative hypothesis that risk factor is associated with increased risk. There was no reason to think or test if they may be associated with reduced risk of PD, because these risk factors were chosen specifically because they have been robustly associated with increased PD risk in other populations. Given that the alternative hypothesis was one-sided (positive direction), tests were conducted one-sided. To test each risk factor individually, OR, lower bound (LB) of 95% confidence intervals (CI), and one-sided P values were calculated with no covariate adjustment from logistic regression models using ‘glm’ in R. To assess strength and significance of each risk factor in the presence of, and adjusted for, other risk factors we used conditional analysis, where we included all relevant risk factors in the regression model, as shown in equations (EQ) (1)-(3):

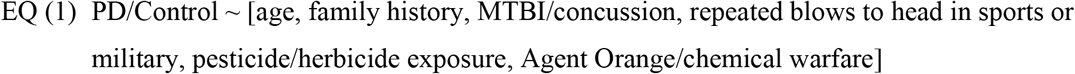

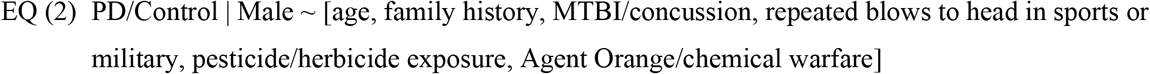

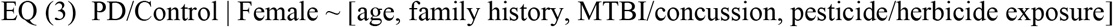

Sex was not included in conditional models due to its collinearity with repeated blows to head, and Agent Orange/chemical warfare. One-sided P value was calculated using ‘glm’ specifying ‘family=“binomial”’ in R. Adjusted odds ratio was calculated using equation ‘exp(x)’ in R where x is a vector of coefficients extracted from ‘glm’ in R. The lower-bound for the one-sided 95% CI were calculated by taking the lower bound of a two-sided 90% Wald CI, using ‘exp(confint.default(x, level=0·9))’ in R where x is a ‘glm’ object from running the conditional models, and level=0·9 specifies the two-sided 90% interval. Analyses were repeated for males and females separately.

Population attributable fraction was calculated using Miettinen formula^29^:

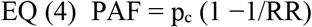

Where p_c_ is prevalence of risk factor in cases and RR is relative risk approximated by OR under the rare disease assumption. To adjust PAF for potential confounding, we used adjusted OR values derived from conditional analysis^30^. For the 95% CI, we used the function ‘AFglm’ from the AF R package on the regression models, specifying ‘case.control=TRUE’, using a sandwich estimator for the variance of PAF^37^.

To assess the fraction of disease attributed to multiple risk factors, we calculated joint PAF (also known as summary PAF)^30^. Prior to calculating joint PAF, in order to meet the assumption of independence and absence of moderating synergy, we tested for pair-wise interaction between the three risk factors associated with PD in males. We used EQ (2) and added an interaction term, as follows:

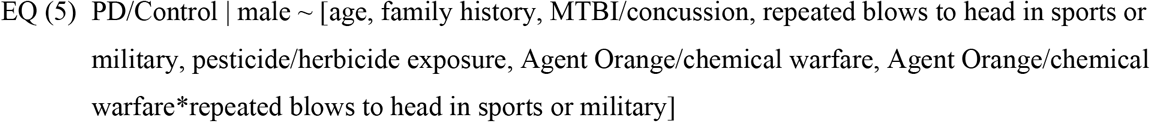

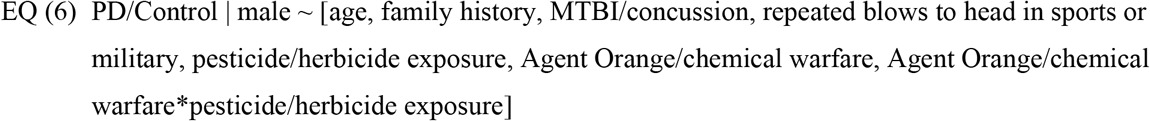

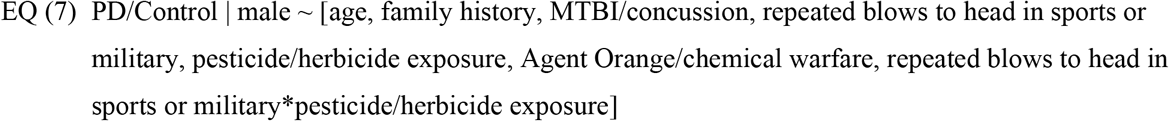

Having established independence and no interaction, we used EQ (8)^30^ to calculate joint PAF

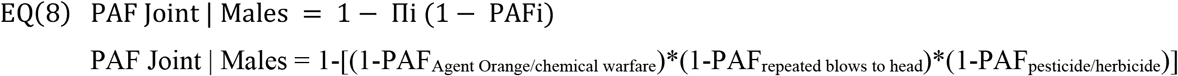

The upper and lower bounds of 95% CI of joint PAF were calculated using EQ (8) with the set of three upper bounds and three lower bounds of the individual adjusted PAFs.

## Supporting information

Supplemental data and Software

## Data Availability

Data will be publicly available without restriction upon peer reviewed publication. The entire dataset at individual level, de-identified, will be provided in Supplementary Material/Data. As per Human Subject Research Privacy considerations, any subject over age 90 years will be denoted as 90 in publicly shared data although true age was used in this analysis. For software used, the URLs and software versions are given in Supplementary Material/Software. The code for reproducing the results can be found on Zenodo [link will be provided in peer reviewed paper].

## Data availability

Data will be publicly available without restriction upon peer reviewed publication. The entire dataset at individual level, de-identified, will be provided in **Supplementary Material/Data**. As per Human Subject Research Privacy considerations, any subject over age 90 years will be denoted as 90 in publicly shared data although true age was used in this analysis. For software used, the URLs and software versions are given in **Supplementary Material/Software**. The code for reproducing the results can be found on Zenodo [link will be provided in peer reviewed paper].

## Acknowledgements

We thank research volunteers for participating in the study. Funding for data collection was provided by a grant from The U.S. Army Medical Research Materiel Command endorsed by the U.S. Army through the Parkinson’s Research Program Investigator-Initiated Research Award under Award No. W81XWH1810508 to Haydeh Payami. Time and effort spent on this project were funded by grants from Aligning Science Across Parkinson’s [ASAP-020527] through the Michael J. Fox Foundation for Parkinson’s Research (MJFF) to Haydeh Payami, NIH/NIEHS 1R01ES032440 to Timothy Sampson, and NIH/NINDS P50 NS108675 to David Standaert. For the purpose of open access, the authors have applied a CC-BY public copyright license to all Author Accepted Manuscripts arising from this submission. The funding agencies had no role in writing of the manuscript or the decision to submit the paper for publication. Opinions, interpretations, conclusions, and recommendations are those of the authors and are not necessarily endorsed by the funding agencies.

## Author contribution

HP conceptualized and designed the study, and oversaw data collection, quality control, and statistical analysis. DGS provided clinic access for patient recruitment and took responsibility for clinical aspects of study. HP, GC and ZDW directly accessed and verified the underlying data. HP, GC, CFM, ZDW processed and analyzed the data. TRS provided expertise on toxicants. HP and TRS wrote the first draft of the manuscript, and all authors reviewed, edited, and approved the final version of the manuscript.

