## Supplemental data and Software for "Population fraction of Parkinson’s disease attributable to preventable risk factors"

**Supplementary data.** Full data used in study.

| Sample ID | Case status | Sex | Age | PD oset age | RBD | MTBI/ concussion | MTBI/ concussion age | Repeated blows to head | Agent orange /chemical warfare exposure | Pesticide/ herbicide exposure | Pesticide/ herbicide exposure duration | State subject born | State current residence | Hispanic or Latino | Race | Jewish ancestry | Family history PD | Family history PD degree | Lost 10 lbs last year | Constipation |
|---|---|---|---|---|---|---|---|---|---|---|---|---|---|---|---|---|---|---|---|---|
| PAF0001 | | | | | | | | | | | | | | | | | | | | |
| PAF0002 | | | | | | | | | | | | | | | | | | | | |
| ... | | | | | | | | | | | | | | | | | | | | |
| ... | | | | | | | | | | | | | | | | | | | | |
| PAF1223 | | | | | | | | | | | | | | | | | | | | |

Data will be populated with peer-reviwed publication.

**Supplementary Material / Software used in the study**

| Software or database name | Version | URL | RRIDs if applicable |
|---|---|---|---|
| PROGENY | 9 | http://www.progenygenetics.com/ | RRID:SCR_006647 |
| Microsoft Excel | 16.0.15601.20526 | https://www.microsoft.com/en-gb/ | RRID:SCR_016137 |
| R | 4.1.3 | https://cran.r-project.org/bin/windows/base/old/4.1.3/ | RRID:SCR_001905 |
| RStudio Connect | 2023.03.0 | https://docs.posit.co/previous-versions/connect/ | RRID:SCR_000432 |
| AF R Package | 0.1.5 | https://cran.r-project.org/package=AF | |
| data.table R Package | 1.14.0 | https://cran.r-project.org/package=data | |
| ggplot2 R Package | 3.4.2 | https://cran.r-project.org/package=ggplot2 | |
| maps R Package | 3.4.1 | https://cran.r-project.org/package=maps | |
| mapsproj R Package | 1.2.11 | https://cran.r-project.org/package=mapsproj | |
| officer R Package | 0.6.1 | https://cran.r-project.org/package=officer | |
| openxlsx R Package | 4.2.3 | https://cran.r-project.org/package=openxlsx | |
| pairwiseCI R Package | 0.1-27 | https://cran.r-project.org/package=pairwiseCI | |
| renv R Package | 0.13.2 | https://cran.r-project.org/package=renv | |
| table1 R Package | 1.4.3 | https://cran.r-project.org/package=table | |
| targets R Package | 0.13.1 | https://cran.r-project.org/package=targets | |
